# Predictive and Causal Analysis of No-Shows for Medical Exams During COVID-19: A Case Study of Breast Imaging in a Nationwide Israeli Health Organization

**DOI:** 10.1101/2021.03.12.21253358

**Authors:** Michal Ozery-Flato, Ora Pinchasov, Miel Dabush-Kasa, Efrat Hexter, Gabriel Chodick, Michal Guindy, Michal Rosen-Zvi

**Affiliations:** IBM Research-Haifa, Haifa, Israel; Assuta Medical Centers, Tel Aviv, Israel; Maccabi Healthcare Services, Tel Aviv, Israel; Tel Aviv University, Tel Aviv, Israel; Ben Gurion University of the Negev, Beer Sheva, Israel; The Hebrew University, Jerusalem, Israel

## Abstract

“No-shows”, defined as missed appointments or late cancellations, is a central problem in healthcare systems. It has appeared to intensify during the COVID-19 pandemic and the nonpharmaceutical interventions, such as closures, taken to slow its spread. No-shows interfere with patients’ continuous care, lead to inefficient utilization of medical resources, and increase healthcare costs. We present a comprehensive analysis of no-shows for breast imaging appointments made during 2020 in a large medical network in Israel. We applied advanced machine learning methods to provide insights into novel and known predictors. Additionally, we employed causal inference methodology to infer the effect of closures on no-shows, after accounting for confounding biases, and demonstrate the superiority of adversarial balancing over inverse probability weighting in correcting these biases. Our results imply that a patient’s perceived risk of cancer and the COVID-19 time-based factors are major predictors. Further, we reveal that closures impact patients over 60, but not patients undergoing advanced diagnostic examinations.

## Introduction

The outbreak of the coronavirus disease 2019 (COVID-19) pandemic in early 2020 led to a unique unprecedented societal shift. Governments implemented a wide variety of nonpharmaceutical interventions (NPIs) aimed at controlling the disease spread and enforcing social isolation. Confinement and school closure as well as entertainment and cultural sector closure were the predominant NPIs taken by governments in 2020^1^. Accordingly, many radiology departments decreased the number of elective imaging examinations to minimize the spread of infection and free up much needed medical resources and staff^2–4^. One specific example is the reduction of breast imaging to 28% of baseline imaging volumes reported by Stanford Health Care during the first months of the pandemic outbreak^3^. Another is the 85% drop in mammography examinations over five days during March 2020^4^ as reported by a large, metropolitan hospital system consisting of six outpatient practices across three New York City boroughs. Many radiology centers in the United States started to gradually reopen these examinations in the summer of 2020^2^. In parallel to the variability in the outpatient radiology practices regarding examination availability, there was also a change in the behavior of patients coming to the exams, showing a distinct rise in no-show rate that seemed to be correlated with the height of the pandemic.

No-shows to medical appointments significantly impact revenue, cost, and the efficient use of resources. Understanding this issue can help us to better manage resources, target interventions to prevent the no-show, and reduce some of the associated revenue loss and cost. Predicting patient no-shows and identifying the major individual attributes associated with no-show have been the key approach to understanding this phenomenon^5,6^. Patient cohorts with retrospective data derived from electronic health records (EHRs) have been leveraged to train prediction models, with logistic regression being the most widely-used model^5,6^. Specifically, among the different modalities used in radiology examinations, mammography (MG) and CT were identified as having the highest no-show rates^7^. Among the different predictors, age and scheduling lead time were often identified as highly associated with not showing up.

In recent years, scalable methods were developed to better understand different phenomena by applying causal inference to patient cohorts^8^. Causal inference can help quantify the magnitude of the effect of specific interventions on no-shows. In the context of the pandemic and the NPIs imposed by governments, it is important to assess the effect of NPIs on the no-shows. Following the common design of causal inference studies, we analyze two groups in the data: appointments that took place when NPIs were not imposed and appointments that took place during the time of imposed NPIs. Inverse Probability Weighting (IPW) is a method frequently used to correct for unequal confounders distribution across the groups. It is aimed at eliminating biases in the data and enabling an inference of the average effect at the population level^9^. However, IPW is not guaranteed to successfully remove biases. A recently developed method called Adversarial Balancing (AdvBal)^10^, based on generative adversarial networks (GANs), is better posed to address covariates biases in high-dimensional data and generate weights that are closer to uniform. We tested and compared IPW and AdvBal to estimate the average effect of the NPIs on the no-show rate.

In this study we analyze the data of patients coming from the largest private network of hospitals in Israel, which performs 30% of the screening mammography exams in the country. Excluding several days, which were dismissed from our no-shows analysis, this network did not apply any policy to decrease the number of imaging exams during the COVID-19 pandemic, but rather adjusted service to changes in the demand. We present a comprehensive study of no-shows during COVID-19, taking into account time-based coronavirus factors such as % confirmed cases, reproduction rate, NPIs, and changes in population mobility, which are derived from multiple data resources. See Figure 1 for the trend of selected time-based statistics over the year 2020. The correlations between changes in the growth of COVID-19 morbidity, population mobility, and the ratio of show-to-no-show are easily noticed. To the best of our knowledge, this is the first study to combine pre-pandemic known predictors of no-show with pandemic associated predictors. Our study is also unique in the causal inference analysis that we perform to assess the average effect of NPIs on no-show in the entire study population, and in subgroups of interest.

**Figure 1.**
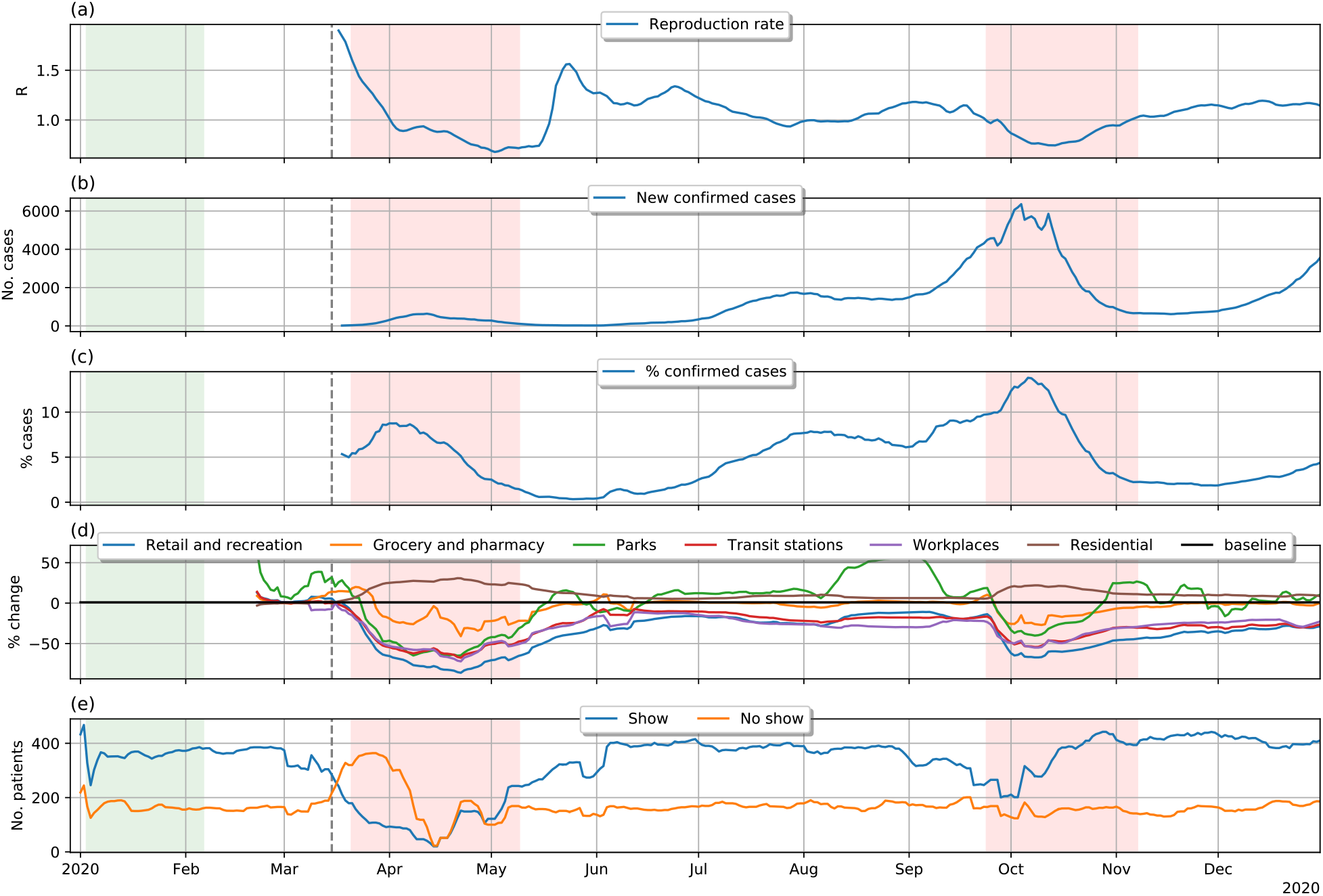
Coronavirus related trends. The dashed lines mark the beginning of the COVID-19 period analyzed in this study; the green rectangle denote a pre-COVID-19 period used as a reference; red rectangles denote school closures.

## Methods

Our study analyzes no-show appointments over the course of 2020. We refer to the time period from January 3 till February 6, 2020 as the *Pre-COVID-19 period*, and to March 15 till December 31, 2020 as the *COVID-19 period*. The Pre-COVID-19 period was defined in a manner similar to the baseline period used in Google Community Mobility Reports^11^. We focused on the COVID-19 period and used the Pre-COVID-19 period as a reference for its comparison. The inspection of NPI data revealed large correlations in these data, as expected. Therefore, we decided to include in our analysis a single NPI, school closure, which is more easily defined, and serves as an indication for other NPIs. Similar to previous studies^12,13^, we consider late cancellations as no-shows since they do not allow the scheduling system to set an appointment for a new patient. We randomly partitioned the patients in this study into train (56%), validation (14%), and test (30%). We trained our prediction models on the train dataset and evaluated them on the validation dataset during the development process. The train and validation sets were also used for the development of the causal analysis. The test dataset was held-out until the model, analysis, and all hypotheses were finalized. Unless specified otherwise, the results presented in this study are from the held-out data.

### Data

This retrospective study was approved by the institutional review board of Assuta Medical Center (0033-20-ASMC), who waived the requirement for patient consent. The appointments dataset was extracted from the EHR datamart of Assuta Medical Centers and included all imaging appointments from January 1, 2020 to December 31, 2020. Our initial cohort included all patients with mammography (MG) and/or breast ultrasound (BUS) appointments during 2020. In addition to the scheduled date, the information on each appointment included the creation and cancellation dates, with the latter filled only for cancelled appointments. Rescheduling an appointment involves cancelling the old appointment and creating a new one, and therefore all dates in an appointment record are assumed non-modifiable. The information on the scheduled exam included: the imaging modality (MG, BUS), the procedure code, a pre-exam urgency indicator, and the site of the exam. The information on the patient included: a (de-identified) patient-ID, gender, age, and town of residence. We received additional data on each patient selected for our study, which included the results of the patient’s last MG, BUS, breast MRI, and breast biopsy exams, if present.

We obtained socio-economic measures and population sizes for localities in Israel from the Central Bureau of Statistics (CBS) reports^14^. We downloaded Israel Transverse Mercator (ITM) coordinates for localities in Israel from the public Israeli Government Databases^15^ and used these data to compute pairwise distances between the patients’ towns and exam sites. Time-based statistics of coronavirus morbidity in Israel, and for specific towns, were downloaded from the COVID-19 Data Repository of the Israeli Ministry of Health^16^. We obtained data on NPIs in Israel from the IBM Worldwide Non-pharmaceutical Interventions Tracker for COVID-19 (WNTRAC)^17^. The data included information on confinements, school closures, public services closures, work restrictions, and more. Finally, we acquired community mobility data in Israel from Google Community Mobility Reports^11^. These reports include the per-day change in movement across six different categories of places, with respect to the median value of the same day of the week during the five-week period January 3 to February 6, 2020.

### No-Show Outcome

We defined a no-show outcome for a pair of (patient, date), if the patient had an appointment for an imaging exam on that date. There were many patients with multiple appointments on the same date, e.g., MG and BUS for breast cancer screening were often scheduled for the same date. We classified a pair of (patient, date) as *show* if the patient attended at least one appointment at that date, and *no-show* if the patient attended none. Our assumption was that partial attendance to appointments is likely not the patient’s choice. We inferred that a site had no service for a certain modality on a specific date, if no corresponding exams were performed on that date. We then refined the no-show definition to exclude (patient, date) pairs for which there was no service for all patient appointments on that date.

### Cohort Selection and Index Date

Our initial cohort included all (patient, date) pairs during the year 2020, where the patient had an ambulatory appointment for an MG or BUS exam on that date. We included only patients aged 18 years or older. We set the index date of each (patient, date) sample to be one-week (7 days) prior to that date, and discarded (patient, date) samples for which all appointments were created at or after the index date. We referred to these discarded samples as “same week schedules” and analyzed them in a separate study, as these were shown to have distinct characteristics. We also excluded (patient, date) samples for which all appointments were cancelled prior to the index date. We refer to such samples as “early cancellations” and assumed the health system can accommodate for these. To summarize, we selected (patient, date) samples with at least one planned appointment by the index date, by excluding samples that had all their appointments cancelled at that date, or not yet created. The process of cohort extraction is depicted in Figure 2.

**Figure 2.**
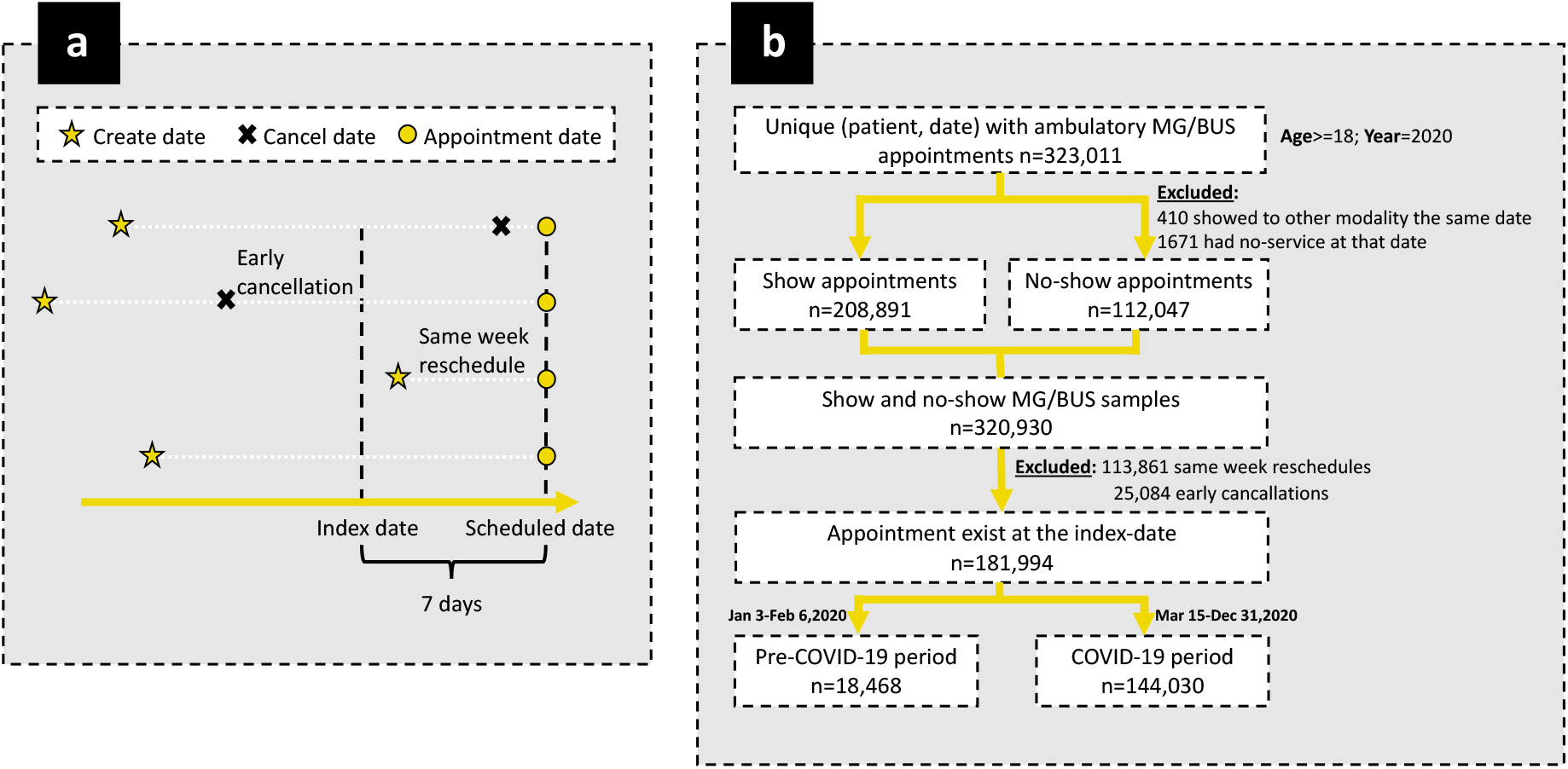
Cohorts’ inclusion/exclusion criteria.

### Features

We used the data from the time period prior to the index date to extract features for our no-show prediction model. The same set of features was used in the analysis of closure effects. We extracted a total of 140 features, using a previously described feature extraction tool^18^. Below we present these features:

#### Patient demographics

age, gender, town socio-economic index-value, town distance from site.

#### Index exams

waiting time (time between the date of the appointment creation and the scheduled date, aka “lead time”), imaging modality, total number of modalities, MG/BUS procedure indicator, total number of MG/BUS procedures, pre-exam urgency indicator, site indicator.

#### Imaging history (3 months)

total number of show dates (all imaging modalities), total number of show dates for the same modality, total number of no-show dates (all modalities), total number of no-show dates for the same modality.

#### Cancer diagnostic phase

we mapped each MG/BUS procedure to a cancer diagnostic phase. Basic and extended breast cancer screening procedures were mapped to phases 1 and 2 respectively; diagnostic exams of patients recalled after screening exams were mapped to phase 3; phases 4 and 5 pertain to biopsy-related procedures, indicating the existence of findings suspected as potentially malignant. The feature “is screening” indicates basic screening (phase 1).

#### Previous MG and BUS exams

has a previous exam indicator, time to last exam, last procedure indicator, last cancer diagnostic phase, last procedure is screening, last BI-RADS score (i.e. the result of the exam), last breast density measure (1-4).

#### Previous breast MRI and biopsy exams

has a previous MRI indicator, time to last MRI, MRI procedure indicator.

#### COVID-19 related measures

\# new confirmed cases, % confirmed cases, change in population mobility for each of the six place categories, school closure indicator, reproduction rate. All these features refer to measures at the country-level. We also extracted town-specific time-based features corresponding to the patient’s town: total number of new confirmed cases in town (per 10K), % confirmed cases in town. All time-based features were computed after the underlying time-series data were smoothed with a moving average of seven days. The COVID-19 reproduction rate was inferred with the EpiEstim^19^ application.

### Predictive Analysis of No-Shows

We trained our no-show prediction model with the gradient boosted trees algorithm, using the XGBoost^20^ package. Hyperparameters were tuned with 5-fold cross validation on the train dataset, via a randomized grid search. We were unable to map the town of residence to the list of towns published by the Israel CBS for 7% of the samples, with a higher rate for no-show samples. Inspecting the unmapped town values revealed that most of these corresponded to either “unknown” or “other” values. We preprocessed the train data and imputed missing values in town-based features, to prevent XGBoost from learning predictive patterns of missing values in these features. The imputation was done with the stochastic “multivariate imputation by chained equations” (MICE) method, with ten iterations, using the “Statsmodels”^21^ package. We relied on the inherent XGBoost imputation mechanism to handle missing values in the remaining features during train, and in all the features during inference. We used TreeExplainer from the Shapley Additive exPlanations (SHAP)^22^ package to estimate the contribution of each feature to our XGBoost model, based on the classic game-theoretic Shapley values. We obtained an agglomerative hierarchical clustering of time-series data using average linkage, and the Euclidean distance between the pairwise Pearson correlations as the distance metric.

### Causal Analysis of Closures

We divided our (patient, date) samples into closure and no-closure groups based on whether there was a school closure on the date. We leveraged the average treatment effect (ATE) to examine the effect of school closure on no-show rate:

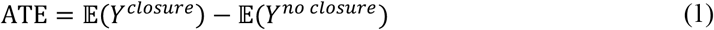

where 𝔼 (*Y*^1^) and 𝔼 (*Y*^0^) are the potential no-show rates for the entire cohort if all samples were during closure and no-closure, respectively. In other words, the ATE is the average effect, at the population level, of moving the entire study population from no-closure to closure. Estimating the ATE from the observed data requires accounting for confounding biases between closure and no-closure groups. For each feature, we measured the bias between the two, possibly weighted, closure and no-closure groups by computing the absolute standardized difference^9^:

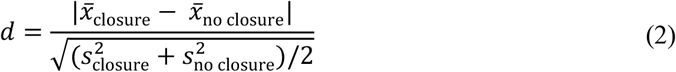

where 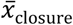 and 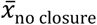 are the feature means in the closure and no-closure groups, respectively, and 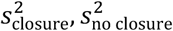 are the corresponding sample variances. A standardized difference that is less than 0.1 is commonly regarded as negligible bias between compared groups^9^. We visually compared the distributions of top-biased features within the closure and no-closure groups. We plotted the smoothed densities of the compared distributions using Gaussian kernel density estimation with a bandwidth of 1.

#### Inverse probability weighting (IPW)^9^

IPW is a longstanding popular method that weights samples based on the inverse of their propensity to be in their assigned group. The propensity is estimated by fitting a model that predicts the assigned group. We used a logistic regression model, which is commonly used for IPW.

#### Adversarial balancing (AdvBal) ^10^

The AdvBal method borrows principles from GANs, to generate sample weights for a source data, such that the resulting weighted data becomes similar to a given target data. In our case, it was aimed at generating weights for the closure (resp., no closure) group, to make it indistinguishable from the entire cohort data. Similar to GANs, it operates by alternately training a classifier to distinguish between the source and target data, and a weights generator that uses exponentiated gradient descent to maximize classification error. We applied AdvBal with a logistic regression classifier. We set the number of iterations in AdvBal to ten.

Prior to applying logistic regression modeling to the data, in both IPW and AdvBal, we imputed missing values in all the features using the MICE method. We used the implementation of IPW and AdvBal from the CausalLib^23^ package.

### Evaluation Measures and Statistical Tests

We measured the accuracy of our prediction models using the area under the receiver-operator curve (AUC). We assessed the statistical association between two binary variables with a chi-squared test, and between a binary variable and a non-binary variable with an unpaired one-sided t-test. These univariate statistical tests were performed after omitting missing values in the tested features. We compared two AUCs that were computed on the same data using the DeLong test^24^. We estimated the standard error of computed effects with 100 bootstrapping iterations, and computed P-values by the normal distribution. We used the validation set for testing various hypotheses, from which we selected 25 that seemed most attainable. We applied the Benjamini-Hochberg^25^ procedure to account for multiple testing, and all the 25 P-values reported in this study were significant at false discovery rate lower than 0.002.

## Results

Table 1 presents selected statistics for the train, validation, and held-out datasets for each of the COVID-19 and Pre-COVID-19 periods. As shown, the train, validation and held-out datasets exhibit similar statistics for both periods. A comparison of the COVID-19 and Pre-COVID-19 periods revealed that the no-show rate significantly increased during COVID-19 (35% vs. 31%, *P*-value = 0.0002, chi-squared test), due to the increase in late cancellations (11% vs. 7%). Note that late cancellations also include late reschedules, that is, reschedules during the week before the appointment. Another prominent difference between the two time periods is the shorter average waiting time during COVID-19 (41 vs 44 days, *P*-value = 2 × 10^−186^, t test), which can be explained by the 7% reduction in the average number of appointments per week. Both “%confirmed cases” and “school closure” showed a significant association with no-shows during COVID-19.

**Table 1.**
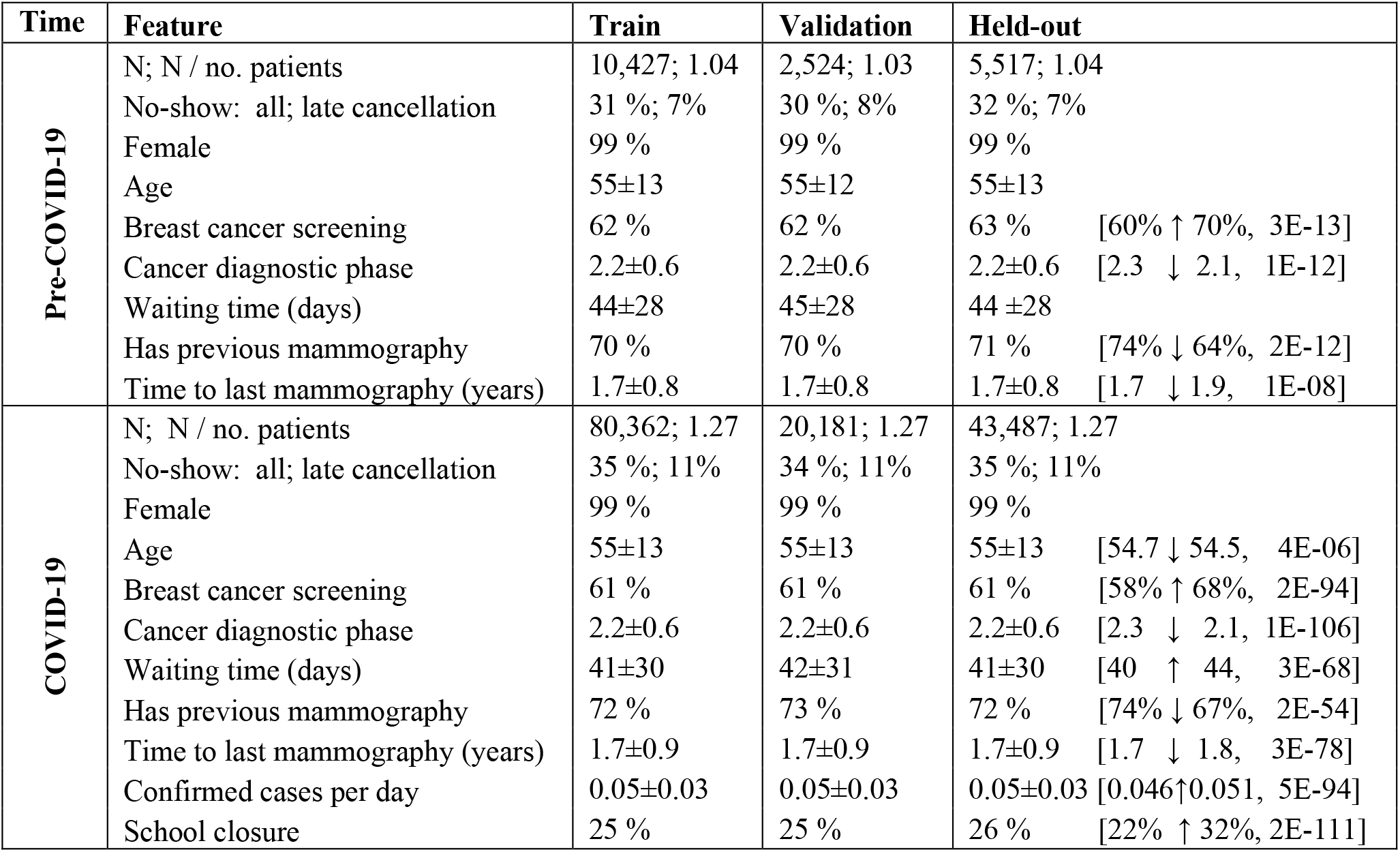
Cohort statistics. For binary variables, we present the proportion (%) and mean ± standard deviation for non-binary features. The association with the no-show outcome (show and no-show statistics, P-value) is presented within squared brackets.

| Time | Feature | Train | Validation | Held-out |
| --- | --- | --- | --- | --- |
| Pre-COVID-19 | N; N / no. patients | 10,427; 1.04 | 2,524; 1.03 | 5,517; 1.04 |
|  | No-show: all; late cancellation | 31 %; 7% | 30 %; 8% | 32 %; 7% |
|  | Female | 99 % | 99 % | 99 % |
| | Age | 55±13 | 55±12 | 55±13 |
| | Breast cancer screening | 62 % | 62 % | 63 % [60% ↑ 70%, 3E-13] |
| | Cancer diagnostic phase | 2.2±0.6 | 2.2±0.6 | 2.2±0.6 [2.3 ↓ 2.1, 1E-12] |
| | Waiting time (days) | 44±28 | 45±28 | 44 ±28 |
| | Has previous mammography | 70 % | 70 % | 71 % [74% ↓ 64%, 2E-12] |
| | Time to last mammography (years) | 1.7±0.8 | 1.7±0.8 | 1.7±0.8 [1.7 ↓ 1.9, 1E-08] |
| COVID-19 | N; N / no. patients | 80,362; 1.27 | 20,181; 1.27 | 43,487; 1.27 |
|  | No-show: all; late cancellation | 35 %; 11% | 34 %; 11% | 35 %; 11% |
|  | Female | 99 % | 99 % | 99 % |
| | Age | 55±13 | 55±13 | 55±13 [54.7 ↓ 54.5, 4E-06] |
| | Breast cancer screening | 61 % | 61 % | 61 % [58% ↑ 68%, 2E-94] |
| | Cancer diagnostic phase | 2.2±0.6 | 2.2±0.6 | 2.2±0.6 [2.3 ↓ 2.1, 1E-106] |
| | Waiting time (days) | 41±30 | 42±31 | 41±30 [40 ↑ 44, 3E-68] |
| | Has previous mammography | 72 % | 73 % | 72 % [74% ↓ 67%, 2E-54] |
| | Time to last mammography (years) | 1.7±0.9 | 1.7±0.9 | 1.7±0.9 [1.7 ↓ 1.8, 3E-78] |
| | Confirmed cases per day | 0.05±0.03 | 0.05±0.03 | 0.05±0.03 [0.046↑0.051, 5E-94] |
| | School closure | 25 % | 25 % | 26 % [22% ↑ 32%, 2E-111] |

### No-Show Predictors During COVID-19

We examined the top-20 contributing features for our XGBoost model that was trained on COVID-19 data with the entire set of 124 extracted features (Figure 3a). The strongest predictor was “cancer diagnostics phase”, an ordinal feature with 5 categories, ranging from basic screening to biopsy related procedures. Additional related features that appear in this list of top-20 were: “last MG cancer diagnostic phase”, and “screening mammography”. The features “last MG BI-RADS score” and “last BUS BI-RADS score” are also related to “cancer diagnostics phase” since BI-RADS scores determine whether patients are recalled for the next diagnostic phase. An inquiry on “procedure BUS 760904”, which was classified as cancer diagnostic phase 2, revealed that it was commonly used when the patient co-scheduled a basic MG screening exam. Thus 6 of the top-20 important features pertain to the patient’s knowledge, or perception, of the a-priori chance of cancer diagnosis. The inspection of “time to last MG”, which was ranked second among the top-20 features, revealed that patients who had mammography exams approximately a year before had the highest tendency to show up on-time to their appointments. However, this tendency decreased as the time from the previous mammography increased.

**Figure 3.**
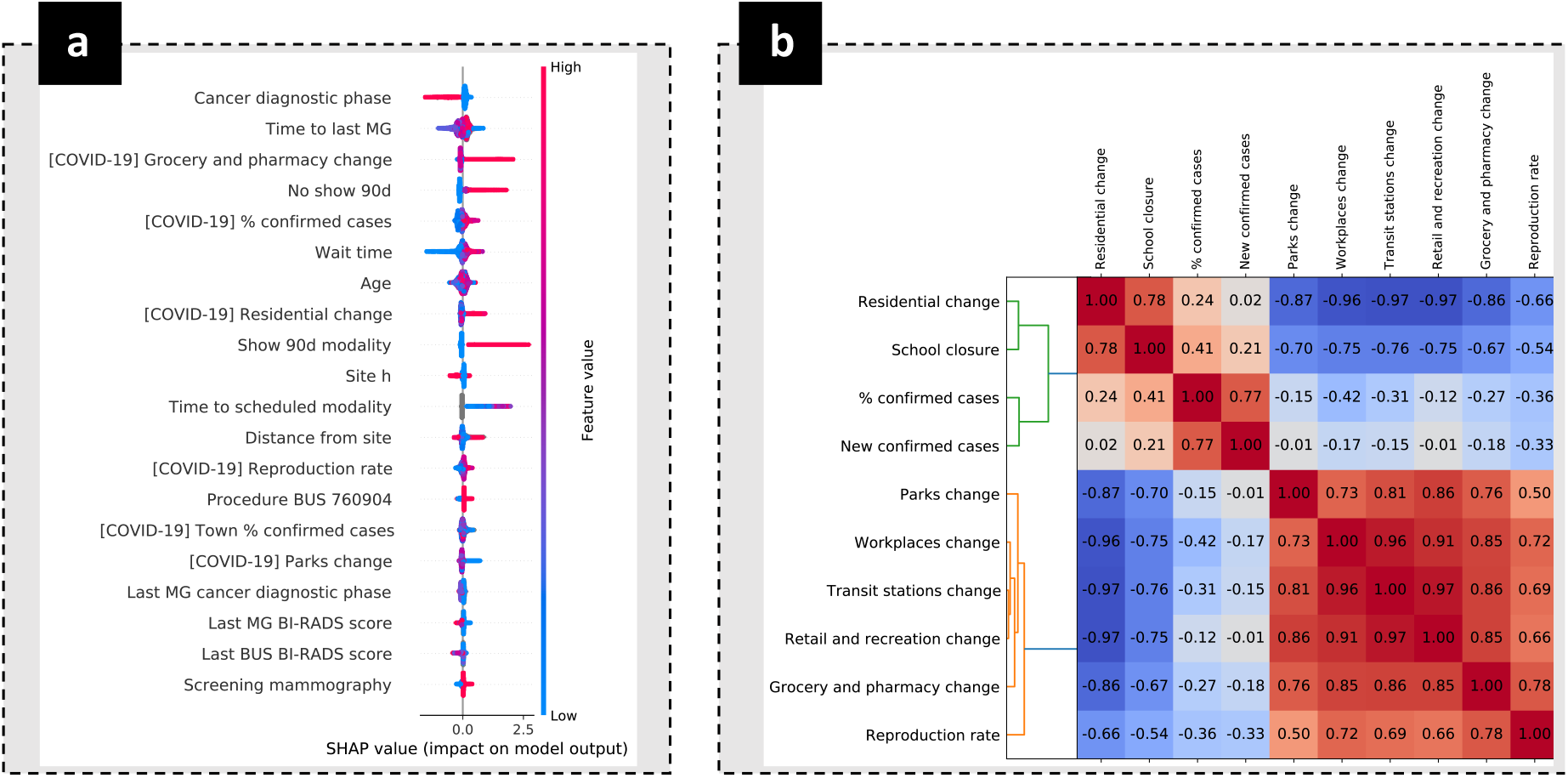
Important predictions and COVID-19 time-series correlations. **a.** 20 top-important features by SHAP analysis. Each point represents a single data sample (i.e., patient at a date). Point color indicates the feature value (red=high, blue=low, gray=missing); X-axis denotes the effect of the feature on predicted no-show score for the sample, which can be positive (increasing no-show predicted score), or negative (decreasing no-show predicted score). **b**. Hierarchical clustering of time-series of COVID-19 related measures.

Six out of the top-20 important features were COVID-19 related factors. The exclusion of these features from our model significantly reduced the AUC from 0.744 to 0.710 (*P*-value = 2 × 10^−90^, DeLong test). Our results imply that when there is a growth in COVID-19 morbidity, patients may be more reluctant to attend MG and BUS exams, as reflected by the positive trend of “% confirmed cases” and “reproduction rate” in Figure 3a. The time-series of COVID-19 measurements largely correlated with each other, and a clustering analysis of these data revealed two clusters that are negatively correlated with each other (see Figure 3b). The correlation of school closure with mobility changes, which were included in list of top-20 important features, may explain the exclusion of the former from that list. The positive association of no-shows with “grocery and pharmacy change” can be explained by the tendency of the latter to increase at the beginning of closures (see Figure 1).

Similar to previous studies^5,7^, we also observed that no-shows were associated with a patient’s history of no-shows (“no show 90d”) and longer waiting time. The distance of the patient’s town from the exam site was also found to be predictive, showing a positive correlation with no-shows for larger distances over 25km. The fact that many of the no-show patients were likely to have their appointments rescheduled was implied by the importance of “show same modality last 90d”, which indicated the patient had recently attended a similar exam, and “time to scheduled modality”, which specified the time to a similar future exam during the year 2020. For the latter, the existence of such an appointment was predictive for no-show, especially when the appointment was further away. Age was found to be predictive as well, but without a clear trend. Finally, following the selection of a site indicator (“site h”) as a top-important feature, we assessed the importance of site indicators by excluding them from our model, which led to a significant decrease in the AUC (0.738 vs. 0.744, *P*-value = 2 × 10^−13^ DeLong test).

### Estimated School Closure Effect on No-Shows

We analyzed the average effect of school closures on no-show rate, after correcting for observed confounding biases between the two time periods. As mentioned above, school closure was selected as a representative of a larger set of highly correlated NPIs and its estimated effect corresponds to the entire set of co-occurring NPIs. We estimated the average effect of school closure in our entire study population, as well as for subgroups defined by (i) age and (ii) cancer diagnostic phase. Our set of potential confounders included all extracted features, excluding school closure itself, and the reproduction rate and mobility features, which were highly correlated with the school closure indicator (See Figure 3b). We tested and compared two causal inference methods for correcting observed biases: IPW and AdvBal. We used a logistic regression model in both methods.

#### Comparison of IPW and AdvBal

In all our analyses, AdvBal substantially diminished the standardized difference of all the confounders (d ≤ 0.02). Reducing the number of the iterations in AdvBal to 5 instead of 10 also successfully reduced biases below the required threshold of 0.1 (d ≤ 0.08). On the other hand, IPW failed to reduce major imbalances, as shown in Figure 4; it even introduced much larger biases into confounders that had negligible biases in the original (unweighted) data.

**Figure 4.**
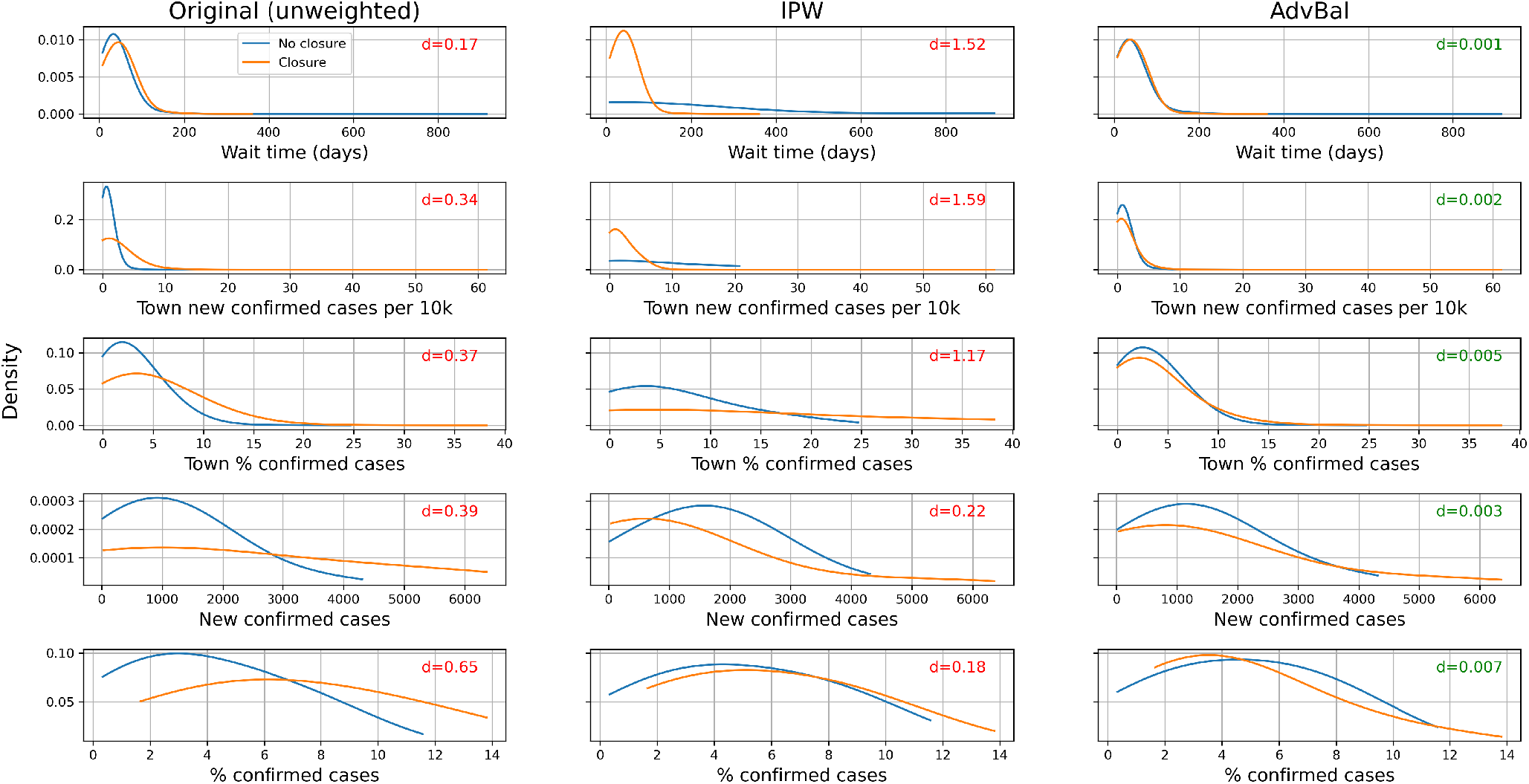
Distribution of top-biased confounders during closure and no-closure periods. Plots depict smoothed estimated densities for the 5 biased features in the original data (left column, d>0.1), and after reweighting the data with IPW (middle column, d>0.1) and AdvBal (right column, d≤ 0.007). The standardized difference d (see Methods section), which measures the bias between the closure and no-closure groups, appears at the upper right corner of each subplot. Green and red colors indicate negligible (≤ 0.1) and non-negligible (>0.1) values for d, respectively.

#### Estimated effects

Table 2 presents the uncorrected and corrected effects of school closure on no-show rates, in the entire study population (“All”) and for subgroups by age and cancer diagnostic phase. As shown, the estimated effect practically vanished in the subgroup of patients with advanced cancer diagnostic exams (phases 3-5), and the remaining estimated effects also largely dropped, with a decrease of 60% to 80% of the original (uncorrected) effects. As an example, the estimated average effect in the entire study population dropped from 12% to 3% after accounting for the observed confounding biases. Among tested subgroups, patients at age 60+ were most affected by closures, with the estimated potential no-show rate lifting from 31% to 37%.

**Table 2.** Uncorrected and corrected average closure effects on no-show rate. Estimated effects are presented with the estimated %no-shows for no-closure and closure groups. Negligible (d<0.1) and non-negligible(d>0.1) standardized difference values are colored in green and red, respectively.

| Group | N | % of cohort | %no-show | % in closures | Uncorrected |  | Corrected (AdvBal) |  |
| --- | --- | --- | --- | --- | --- | --- | --- | --- |
|  |  |  |  |  | Estimated effect | Max. bias (d) | Estimated effect | Max. bias (d) |
| All | 43,487 | 100% | 35% | 26% | 12%: 32%↑43% | 0.65 | 3%: 33%↑36% | 0.01 |
| **Cancer diagnostic phase** |  |  |  |  |  |  |  |  |
| 1-2 | 35,263 | 81% | 38% | 26% | 13%: 34%↑47% | 0.65 | 4%: 36%↑40% | 0.02 |
| 3-5 | 8,224 | 19% | 21% | 24% | 6%: 20%↑26% | 0.67 | 0%: 21%→21% | 0.02 |
| **Age (years)** |  |  |  |  |  |  |  |  |
| <60 | 27,927 | 64% | 35% | 26% | 10%: 32%↑42% | 0.65 | 2%: 34% ↑ 36% | 0.02 |
| ≥60 | 15,560 | 36% | 34% | 25% | 15%: 30%↑46% | 0.66 | 6%: 31% ↑ 37% | 0.01 |

## Discussion

This paper presents a comprehensive analysis of no-shows, defined as missed appointments or late cancellations, during the COVID-19 pandemic, systematically verifying previously made hypotheses^3^. Measures of COVID-19 growth, such as “%confirmed cases”, “new confirmed cases”, and the reproduction rate, are often published in the Israeli media. Our results suggest these representations of the perceived severity of the pandemic play a major role in a patient’s decision of whether to attend a medical appointment. During 2020, there were two periods of school closures in Israel, in which additional non-pharmaceutical interventions were applied, such as confinements and bans of mass gatherings. These closures covered 25% of the appointments in our data during the COVID-19 period (see Table 1). We demonstrated that the observed large effect of school closures on no-show rate dramatically decreased by 60% to 80% when accounting for biases in COVID-19 growth measures. Moreover, we show that closure effect was magnified for patients over 60 years, presumably due to their elevated risk for coronavirus complications. Conversely, no effect was observed for patients in advanced cancer diagnostic phases. The adjustment for the observed biases was enabled by the adversarial balancing^10^ method, after the application of the popular IPW method failed to balance these biases. Adversarial balancing harnessed the power of GANs to generate balancing weights that made the multivariate distributions of the confounders in the closure and non-closure groups less diverged. In this study, we applied adversarial balancing with a simple logistic regression classifier, but stronger deep learning classifiers can be used to increase its power.

Our results are in agreement with existing literature on no-shows^5,7^: patient’s historical behavior, wait time (aka “lead time”), distance from the exam site, and age were found to be predictive for no-shows. Age was also found to be predictive for no-shows but with mixed effects due to several reasons. First, breast screening in Israel is recommended at ages 50 to 74, and hence women under 40 are more likely to attend MG/BUS exams due to some indication, or suspicion, for elevated risk of breast cancer. In addition, in the screening population (diagnostic phase 1), the cancer risk is known to increase with age; however, patients at age 60+ in our data were at risk for coronavirus complications, which may have caused these patients to refrain from attending medical exams. We also demonstrated that including per-site indicators in the model can significantly improve the model accuracy, as it allows the model to identify predictive patterns that are site-specific. Finally, an important finding of our study is the inclusion of the cancer diagnostic phase as a predictor in our model. This predictor, which may reflect cancer risk as perceived by the patient, was ranked as most influential in our model. This discovery demonstrates that the patient’s comprehension of the health risks is important for reducing no-shows.

## Conclusion

We employed state-of-the-art machine learning and causal inference methods to study no-shows in the data of more than 160,000 screening and diagnostic breast imaging appointments made during 2020. Our results suggest that a patient’s perceived risk for breast cancer, as implied by the diagnostic phase, is the most influential factor in their decision to attend an imaging appointment. Additional novel predictors correspond to time-based factors indicating the growth of COVID-19 morbidity. After adjusting for confounding biases with the AdvBal method, the effect of closures on the no-show rate largely diminished. This was especially noted in patients in advanced diagnostic phases, who seemed to maintain their lower rate of no-shows. On the other hand, the impact of closures was most pronounced in patients at age 60+, presumably due to their higher risk for coronavirus complications. Our study further demonstrates the need for personalized medicine to target additional women at higher risk for breast cancer within subpopulations associated with increased no-show rates. In a future study, we intend to leverage richer clinical data to estimate the coronavirus and cancer risks more accurately and analyze them in the context of no-shows.

## Data Availability

The retrospective data was extracted from the EHR datamart of Assuta Medical Centers and cannot be shared. The other datasets that were analysed in this study are public and their links appear in the manuscript.

https://www.google.com/covid19/mobility/

https://www.cbs.gov.il/en/mediarelease/Pages/2020/Characterization-Classification-Geographical%20Unitsby-%20Socio-Economic-Level-Population%202017.aspx

https://data.gov.il/dataset/828

https://data.gov.il/dataset/covid-19

## Acknowledgements

This study was partially supported by the Israel National Institute for Health Policy Research.

